# Machine learning aided multiscale modelling of the HIV-1 infection in the presence of NRTI therapy

**DOI:** 10.1101/2022.06.14.22276382

**Authors:** Huseyin Tunc, Murat Sari, Seyfullah Kotil

**Affiliations:** Department of Biostatistics and Medical Informatics, School of Medicine, Bahcesehir University, Istanbul, Turkey; Department of Mathematics, Yildiz Technical University, Istanbul, Turkey; Department of Biophysics, Bahcesehir University Medical School, Istanbul, Turkey

## Abstract

The Human Immunodeficiency Virus (HIV) is one of the most common chronic infectious diseases of humans. Increasing the expected lifetime of the patients depends on the use of optimal antiretroviral therapies. The emergence of the drug-resistant strains may decrease the effects of treatments and lead to Acquired Immune Deficiency Syndrome (AIDS) even if the existence of antiretroviral therapy. Investigation of the genotype-phenotype relations is a crucial process to optimize the therapy protocols of the patients. Here we propose a mathematical modelling framework to address the effect of initial strains, initiation timing and adherence levels of nucleotide reverse transcriptase inhibitors (NRTI) on the emergence of a possible AIDS phase. For the first time, we have combined the existing Stanford HIV drug resistance data with a multi-strain within-host ordinary differential equation (ODE) model to track the dynamics of most common NRTI resistant strains. Regardless of the drug choice, the late initiation and poor adherence levels to the NRTI therapy increase the probability of the emergence of the AIDS phase. Overall, the 3TC, D4T-AZT and TDF-D4T drug combinations provide higher success rates. The results are in line with genotype-phenotype data and pharmacokinetic parameters of the NRTI inhibitors, but we show the heavy influence of neighbour viral strains of the initial ones has a considerable effect on the success/failure rates. Improving multiscale models can contribute to understanding the disease progression and treatment options.

## INTRODUCTION

Antiretroviral drug resistance is one of the main barriers to therapy success for HIV-positive patients. According to the WHO HIV drug resistance report 2021, 10% of adults without a history of treatment and 40% of those who do are affected by drug-resistant strains (DRS). In addition, 50% of newly diagnosed infants were exposed to the DRS. The DRS can be acquired with nonadherence to the therapy protocols, or patients can directly be infected with DRSs (Blower et al., 2001). Both scenarios yield life-long persistence of DRS and need to be carefully tracked by clinicians by suggesting optimal therapy protocols.

Quantitative evaluation of HIV drug resistance has been carried out with the use of phenosense assays by finding the fold-change of *IC*_50_ values (the amount of concentration to inhibit %50 of virion) between drug-resistant and wild type strains (Zhang et al., 2005; Pham et al., 2018; Feng et al., 2016). Accounting all possible genotype-phenotype relations with such experiments is time-intensive and expensive. On the other hand, data modelling frameworks have been used to construct general mathematical relations between genotype and phenotype information (Steiner et al., 2020; Tarasova et al., 2018; Shah et al., 2020). These mathematical models aim to generalize the given data by encoding the amino acid sequence of target enzymes (Rhee et al., 2010). One of the main contributions of the current study is to investigate how these models can be embedded into a within-host model to answer some critical questions about HIV dynamics.

In the presence/absence of resistant strains and in antiretroviral therapy, various within-host models have been presented to forecast the viral dynamics of the HIV in ordinary differential equation (ODE) forms (Hadjiandreou et al., 2007; Perelson and Nelson, 1999; Dixit and Perelson, 2004; Rong et al., 2007; Sutimin et al., 2017; Wu and Zhao, 2020; Chen et al., 2021). The proposed mathematical models assume the co-existence of susceptible and resistant strains and generally explore the response of ART. Additionally, the effect of drug adherence on the virologic failure of ARTs (Rosenbloom et al., 2012), time-dependent drug efficiencies on ART response (Rong et al., 2007; Vaidya and Rong, 2017), competition between susceptible and resistant strains on the viral dynamics (Ball et al., 2007; Lythgoe et al., 2013) and latently infected CD4+ T cell reservoirs on the evolution of strains (Doekes et al., 2017) have been investigated through within-host models. The current study addresses similar questions with a novel multiscale model fed by the Stanford HIV Drug Resistance data and machine learning models. A detailed review about the existing within-host and between-host multiscale mathematical models for the HIV dynamics can be found in the literature Dorratoltaj et al. (2017).

Here we have the first time combined the experimental drug resistance data of nucleotide-reverse transcriptase inhibitors (NRTI) from the Stanford HIV drug resistance database with a within-host model of HIV infection to observe the dynamics of the viral strains under different scenarios. We aim to investigate the effect of initiation timing and adherence level of the NRTI therapies on the emergence of the AIDS phase with low CD4+ T cell levels. We propose a novel multiscale and multi-strain within-host model consisting of three major parts: assessment of mono and combined time-dependent NRTI therapy effect by considering within blood properties of each inhibitor, assessment of *IC*_50_ values of each NRTI in the presence of each strain with machine learning models and construction of multi-strain within-host ODE based model including CD4+ T cells and macrophage cells for primary targets of virions. Our model has enabled us to observe a primary trade-off between timing and adherence to ART on the occurrence of the AIDS phase, which has not been investigated through data-driven mathematical models so far.

## MATERIALS AND METHODS

### Within-host model with wild-type virus

In this part, we have inspired from previous studies of the within-host HIV infection model in the literature (Hadjiandreou et al., 2007; Hernandez-Vargas, 2019). We assume that the primary reservoirs for HIV infection are: CD4+T cells and macrophages denoted by *T*(*t*) and *M*(*t*) (Hernandez-Vargas, 2019). The long-living macrophage cells cause the persistence of virions over the years (Orenstein, 2001; Herbein and Varin, 2010). Macrophage cells contribute to the depletion of healthy CD4 + T cells in advanced HIV infection (Crowe, 1995). Within host modelling of the HIV infection without considering the macrophage reservoirs yielded less realistic outcomes, such as the models never resulting in the AIDS phase (Rong et al., 2007). We denote the HIV infected CD4+T cells and macrophages by *T*∗(*t*) and *M*∗(*t*). Lastly, the number of free wild-type virions in the host is denoted by the function *V* (*t*). By considering model assumptions like homeostatic cell proliferation terms (*s*_*T*_, *s*_*M*_), bilinear incidence terms (*k*_*T*_ *TV, k*_*M*_*TM*), natural deaths of cells and virions (*δ*_*T*_ *T, δ*_*M*_*M, δ*_*T*_∗ *T* ∗, *δ*_*M*_∗ *M*∗, *δ*_*V*_*V*), viral replication terms (*p*_*T*_ *T* ∗, *p*_*M*_*M*∗) and the Michaelis-Menten type proliferation terms 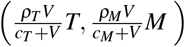, we express the one strain within-host model with the following system of ODEs (Hernandez-Vargas, 2019)

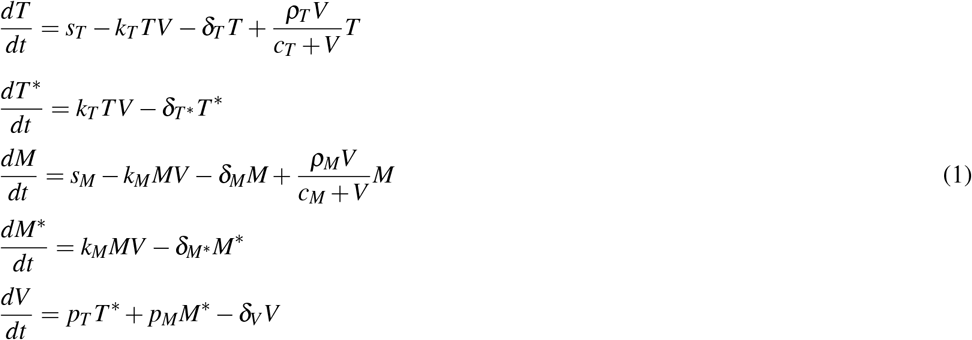

where initial conditions are considered as 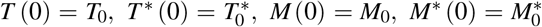 and *V* (0) = *V*_0_. Parameter values of one strain within-host model (1) with the corresponding references can be seen in 1. In the following part, we will extend model (1) to include multi strains (both susceptible and resistant) and NRTI therapy.

### Multi strain within-host model with NRTI therapy

The HAART therapy includes at least one of the NRTIs that aim to block activation of the reverse transcriptase enzyme. Effective therapy of HIV positive patients with NRTIs saves millions of lives worldwide (Tressler and Godfrey, 2012). However, error-prone structure of the HIV replication yields resistant strains over the years and these strains are known to be a primary barrier to preventing AIDS (Kuritzkes, 2011). Our multiscale within-host model includes three main steps: constructing machine learning models to generalize isolate-fold change data for NRTIs, a model for dealing with NRTI action in blood and cells, and finally, a within-host model with multi strains and NRTI therapy.

#### An artificial neural network model for isolate-fold change relation

There exists various genotype-phenotype experiment data, including the fold change values of *IC*_50_ (the required drug concentration to inhibit %50 of virions) for various reverse transcriptase inhibitors in the presence of susceptible and resistant isolates (Rhee et al., 2006). The most used genotype-phenotype data is the Stanford HIV drug resistance database (https://hivdb.stanford.edu/). We use filtered genotype-phenotype data of reverse transcriptase inhibitors available in this database and are widely used for various machine learning algorithms (Amamuddy et al., 2017; Masso and Vaisman, 2013). By regulating the data for each NRTI, 1224 unique mutations have been observed for the reverse transcriptase enzyme. In this filtered dataset, 1662 isolates for epivir (3TC), 1597 isolates for abacavir (ABC), 1683 isolates for zidovudine (AZT), 1693 isolates for stavudin (D4T), 1693 isolates for didanosine (DDI) and 1354 isolates for tenofovir (TDF) have been analyzed for NRTI susceptibility. The dataset includes 1206, 1136, 1220, 1223,1223 and 1119 unique mutations for 3TC, ABC, AZT, D4T, DDI and TDF, respectively.

We apply the binary barcoding technique (Rhee et al., 2010) to represent the isolates that occurred in the dataset. Hence, 1224-dimensional input vectors of 0s and 1s are created by considering the existence of unique mutations in the isolates. Let us denote our complete mutation set as *X* = {*x*_1_, *x*_2_, …, *x*_1224}_ where *x*_*i*_ is an NRTI specified mutation pattern. We define the binary representation of isolate *j* as *I*_*j*_ = {*a*_1_, *a*_2_, …, *a*_1224_} with

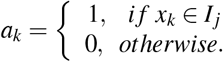

We construct six artificial neural network (ANN) models to predict fold-change values in the presence of any isolates related to each NRTI therapy by using the Machine Learning and Deep Learning toolbox of the MATLAB program. The ANN architectures include 1224-dimensional input, five hidden layer neurons and one output neuron with hyperbolic tangent-sigmoid and linear activation function. The scaled conjugate gradient algorithm with MATLAB built-in function “trainscg” has been used in the training process over GPU. Let us denote our model as a function that maps isolates to the fold changes as

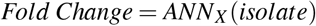

where *X* is a specified inhibitor (*X* ∈ {3TC, ABC, AZT, D4T, DDI, TDF}). In order to overcome possible overfitting, we have implemented an ensemble learning process. For each inhibitor, the 50×100 model has been trained with random training, validation, and test set (80%, 10% and 10%). A model is chosen from every 100 models that yield the minimum mean square error for the test set of the corresponding inhibitor data. Hence, 50 optimal models are chosen among the 5000 models for each NRTI inhibitor, and the final model is calculated as the average of these models.

The prediction performance of six *ANN*_*X*_ (*isolate*) with linear correlation coefficient (R) and mean square error (MSE) values are presented in Figure 1. According to the figure, *ANN*_*X*_ (*isolate*) models yield accurate predictions with high R scores and low MSE scores. Mean MSE value of *ANN*_*X*_ (*isolate*) models have been obtained as 0.0453 with 95% CI [0.0005, 0.0901]. Similarly, the mean *R* value of the models has been calculated as 0.9093 with 95% CI [0.8677, 0.9509].

**Figure 1.**
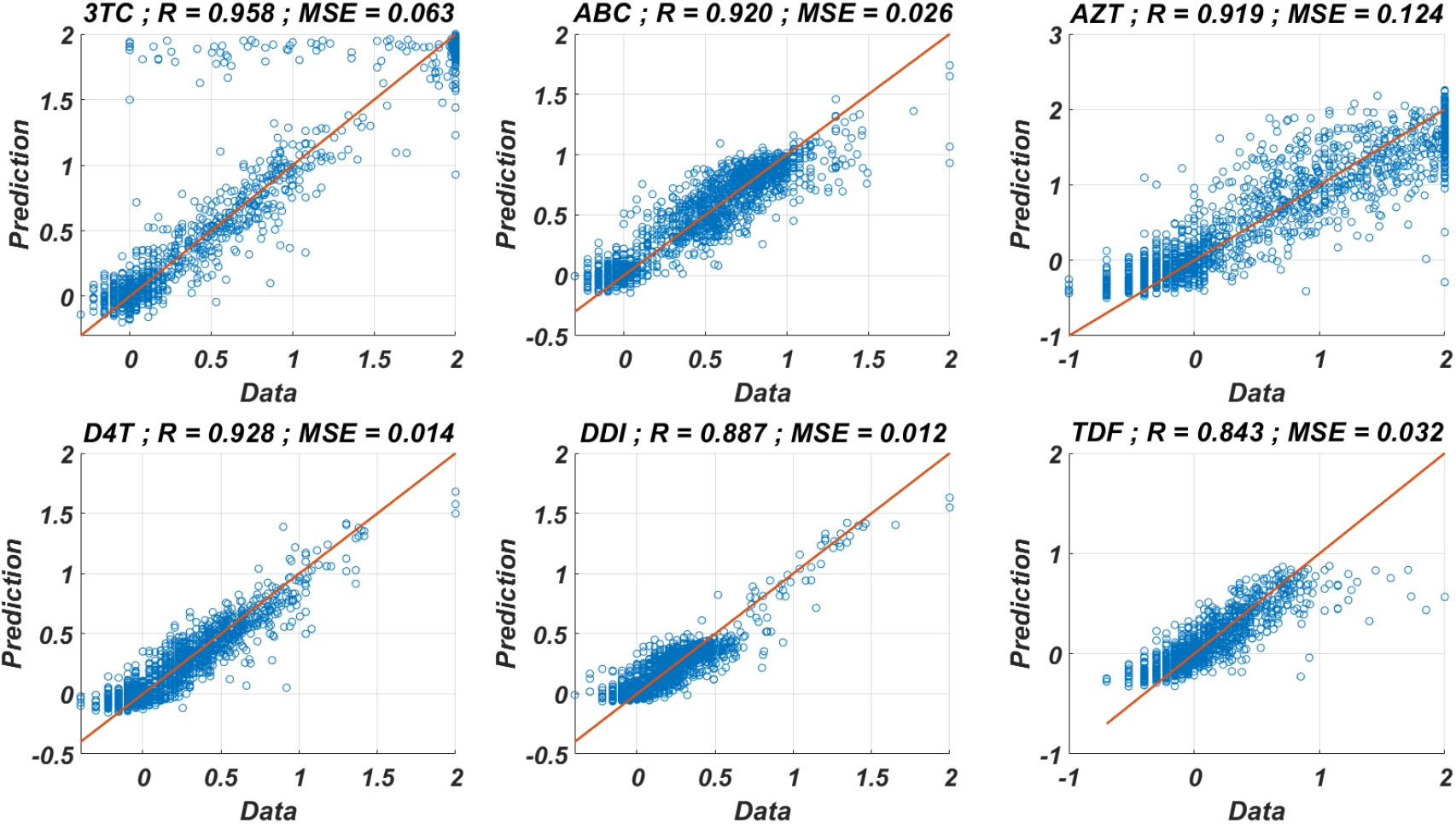
Regression performance of the six ANN models for predicting logarithmic fold change values with corresponding correlation coefficient (R) and mean square error values (MSE).

#### Modelling the time-dependent drug efficacy

Modelling the efficacy of antiretrovirals by using the plasma drug concentrations can be seen in various studies in the literature (Rosenbloom et al., 2012; Rong et al., 2007; Dixit and Perelson, 2004). Here we use the time-dependent drug efficacy model described by Dixit and Perelson (2004) by considering the dynamics of drug concentrations in the blood. Dixit and Perelson (2004) considered the phosphorylated concentration of the tenofovir (TDF) in the cells. Since the time-drug efficiency functions obtained by taking into account blood concentration and phosphorylated within cell concentration of drugs follow a very similar trend, here we assume the blood concentration of the drugs (see Figure 1 of Dixit and Perelson (2004)). Additionally, the non-availability of phosphorylation reaction parameters for the remaining five inhibitors 3TC, ABC, AZT, D4T and DDI have encouraged us to consider the blood concentration of the drugs only.

Let 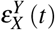 denote the time-dependent efficacy of drug *X* in the presence of strain (isolate) *Y*. The instantaneous efficacy can be approximated by (Dixit and Perelson, 2004)

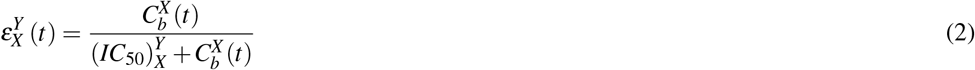

where 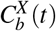 denotes the within blood concentration of drug *X* and 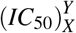 denotes the required concentration of drug *X* to inhibit the 50% of strain *Y*. According to our isolate-fold change ANN model, Eq. (2) can be rewritten as

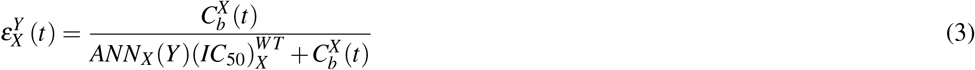

where 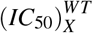 denotes the required concentration of drug *X* to inhibit the %50 wild type virus. Thus, to completely describe 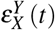, we should clearly express 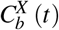. The concentration of a drug in the blood can be expressed as (Dixit and Perelson, 2004)

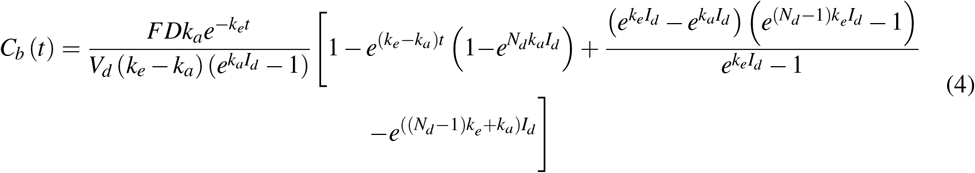

where *F* is the bioavailability of the drug, *D* is the mass of the drug administered in one dose, *I*_*d*_ is the dosing interval, *N*_*d*_ is the number of doses up until time *t, V*_*d*_ is the volume of distribution, *k*_*a*_ and *k*_*e*_ are pharmacokinetic parameters. The drug-specific parameters *k*_*a*_, *k*_*e*_, *D, I*_*d*_ and *F* occurred in Eq. (4) and *IC*_50_ values for 3TC, ABC, AZT, D4T, DDI and TDF according to the equations given by Dixit and Perelson (2004) are evaluated and presented in 1. The detailed explanations of the derivation of these parameters are given in the Supplementary Article and Table S1.

#### A multi-strain within-host model

This part of the study combines all our investigations in a unique multi-strain within-host model. To reduce the cost of the simulations, we assume the main NRTI related mutations 115F, 151M, 184I, 184V, 210W, 215F, 215Y, 41L, 65N, 65R, 67N, 69D, 70E, 70G, 70R, 74I and 74V according to the study of Rhee et al. (2005). These 17 mutations yield 131,071 strains with single, double, triple etc. mutations. Thus, by considering wild-type and mutant strains, we have total *N* = 131, 072 strains. Our multi-strain within-host model with time-dependent NRTI therapy can be derived from one strain model (1) as follows

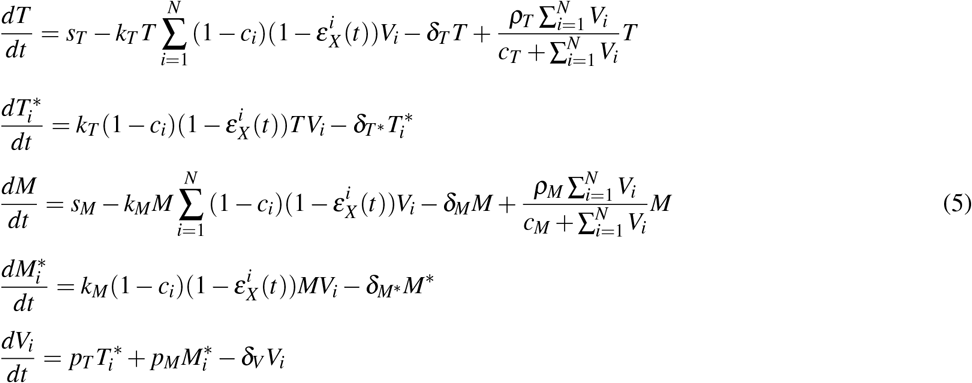

where 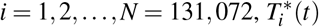 and 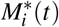 denote the number of CD4 + T cells and macrophage cells infected by strain *i* and *V*_*i*_(*t*) represents the number of virions having *i th* genotype. In the multi-strain within-host model (5), 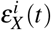 denotes the time-dependent efficacy of the inhibitor *X* on the strain *i* and 0 ≤*c*_*i*_≤ 1 represents the fitness costs of mutant strains with *c*_1_ = 1 for the wild type of strain. The lack of enough experimental results on these fitness values compelled us to use the mean fitness cost values of mutations 41L, 67N, 70R, 184V, 210W, 215D, 215S and 219Q estimated by Kühnert et al. (2018) as 0.2232, 0.3181, 0.3863, 0.5899, 0.3091, 0.0981, 0.1664 and 0.3207, respectively. According to these data, we assume that *c*_*i*_ = 0.3015 for mutant strains *i* ≥ 2. A schematic illustration of the multi-strain within-host model (5) is given in Figure 2.

**Figure 2.**
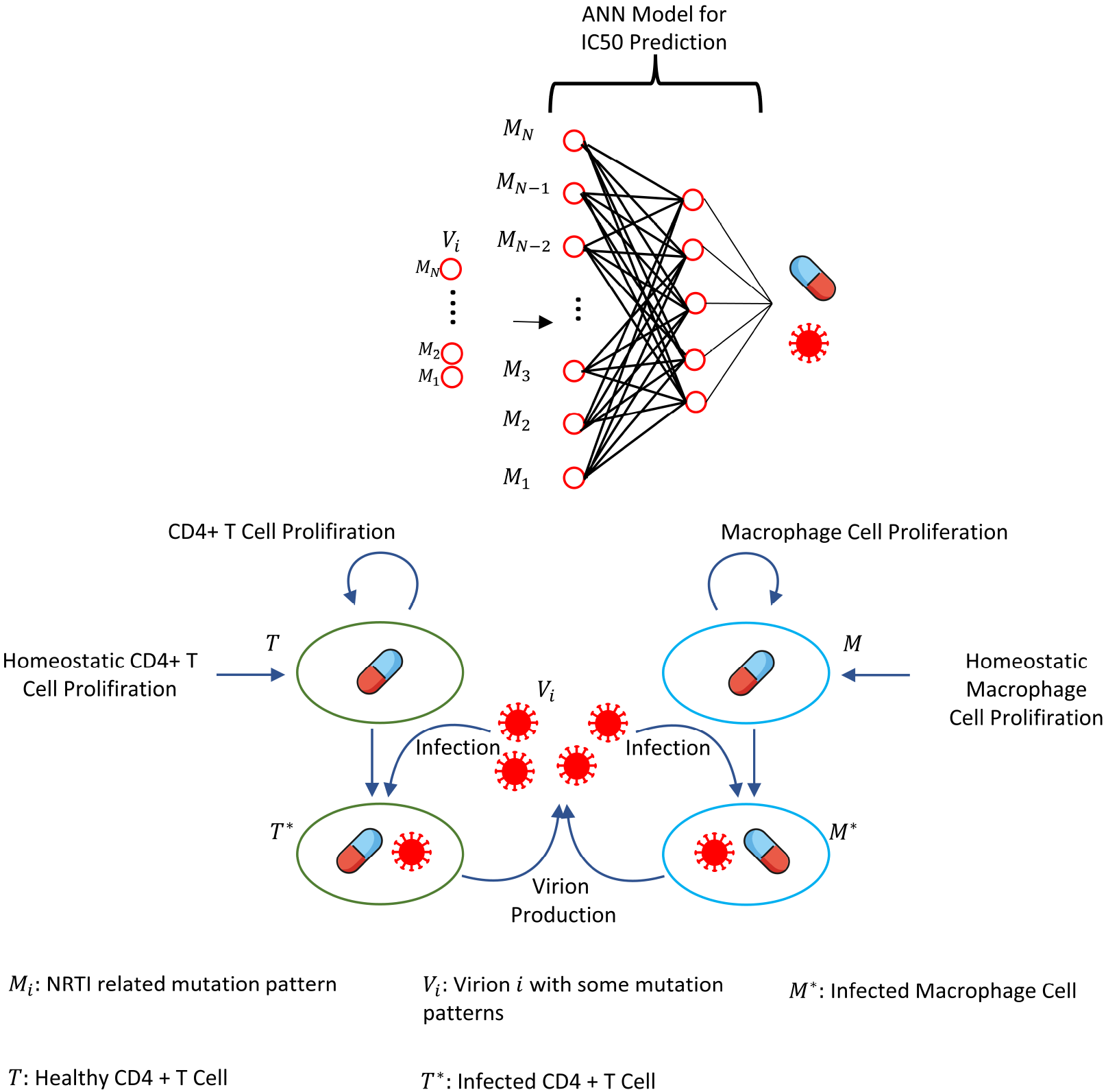
Illustration of the core parts of multi-strain within-host model (5) with NRTI therapy. Model (5) assumes the healthy CD4 + T cells (*T* (*t*)) and macrophage cells (*M* (*t*)) as the main targets of the viral strains (*V*_*i*_ (*t*)). *T*(*t*) and *M*(*t*) increases with both homeostatic cell proliferations and cell proliferations due to the increasing viral load. Viral strains infect both CD4 + T cells and macrophage cells and then those healthy cells become infected CD4 + T cells 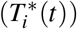 and macrophage cells 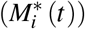. 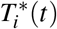 and 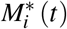 compartmentals produce mature viral strains *V*_*i*_(*t*) with some constant rates. All compartmentals have natural death or clearence with some constant rates. NRTIs block the infection mechanism of the viral strains to the healthy cells. Efficiencies of the NRTIs are estimated through pharmacokinetic equation (3) and the pre-trained artificial neural network models that map the genotype data to fold-change values of the IC50s with respect to the wild type virion.

To realistically model the effect of mutations, we do not explicitly include the mutation matrix in the ODE system (5); instead, we consider the transition between strains by mutations at the end of each time step by generating Poisson random numbers (Rosenbloom et al., 2012). Let us assume time step *n* (*t* = *n day*), 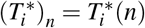 and 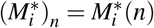. The mutation matrix of our system is denoted by *MT* and defined as

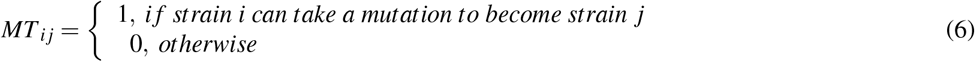

For infected CD4 T cells 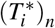 and infected macrophage cells 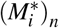, we compute the number of new infected ones in one day period as 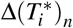 and 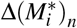 without taking into account the death of these newly infected cells. For each *i* = 1, 2, …, *N, poissrnd* 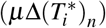 and *poissrnd* 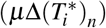 number of infected cells are randomly transmitted from strain *i* to strain *j* according to the mutation matrix *MT*_*ij*_ where *poissrnd*(*x*) function generates the Poisson random number with mean *x* and the mutation rate *µ* = 10^−5^. Thus, this procedure more realistically models the existence of mutations than explicitly embedding the mutation matrix *MT*_*ij*_ to multi-strain within-host model (5). Parameter values of models (1) and (5) are presented with their references in Table 2.

Model (5) can also include dual therapy of NRTIs, *X* and *Y* by modifying the therapy-related time-dependent infection coefficients for CD4 + T cells and macrophage cells 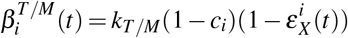 with the use of Bliss independence of drug actions as (Jilek et al., 2012)

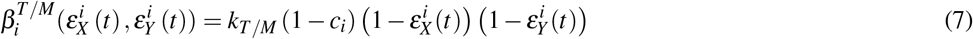

or Loewe additivity of drug actions (Jilek et al., 2012)

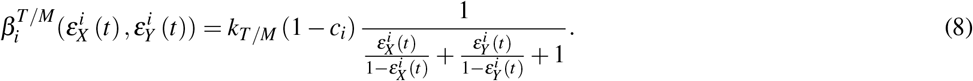

The Bliss independence assumes independent actions of combined drugs, and the Loewe additivity assumes the competition for the same binding site. According to (Jilek et al., 2012), all combinations except AZT-D4T and DDI-TDF obey the Bliss independence rule, and the two combinations obey the Loewe additivity rule. Note that, since we assume *k*_*M*_ = *k*_*T*_ / 1000 according to the Hernandez-Vargas (2019) (see Table 2), 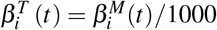, and we prefer to use the notation *β*_*i*_ for 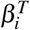 throughout the following parts. Whenever *β*_*i*_ values are quantitatively mentioned in the results section, these values correspond to 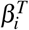.

## RESULTS

This section provides the simulation results of the multi-strain within host model (5), starting with various viral strains. The effects of adherence levels and initiation timing of NRTI therapies on the progression of the AIDS phase are investigated. This section includes four subsections in which we propose the statistics of the infection rates, details of model simulations, the quantitative measure for the therapy success and the simulation results for various cases.

### Statistics of Infection Rates

Before running the simulations to observe the failure/success distribution of each NRTI combination, we may predict the best possible therapy protocol through our pre-trained machine learning model and the pharmacokinetic properties of the inhibitors. Obviously, as we infer from our model (5) and drug-specific time-dependent infection rate 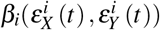 (7)-(8), each viral strain has its infection rate and aims to be dominant by infecting more healthy cells. Since evaluation of 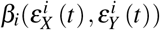 is straightforward through Eqs. (7)-(8) and (3), we can have some priori estimates for the selection of the best therapy protocol. Distribution of 131,071 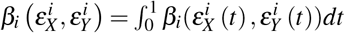 values in the presence of 21 different mono and dual NRTI therapies are illustrated in Figure 3. Descriptive statistic values of 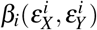 values for all combinations are presented in Table 3.

**Figure 3.**
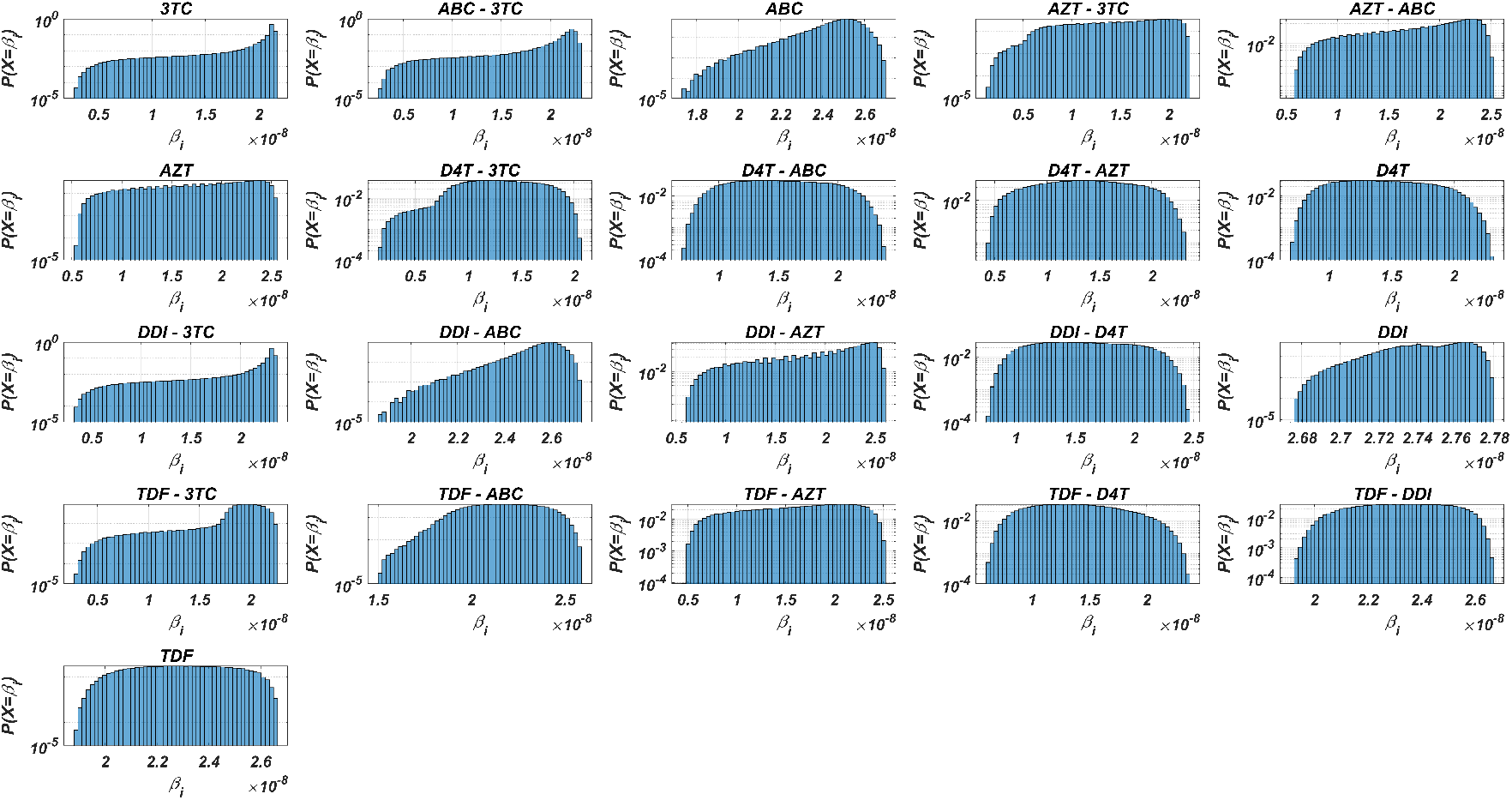
Probability distributions of infection rate (*β*_*i*_) values of various viral strains in the presence of NRTI therapy combinations.

Figure 3 and 3 show that the probability distributions are almost uniform and 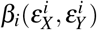 values have considerable diversity and standard deviations among the viral strains. Hence, this observation means that even having point mutations may change the infection rates considerably and thus may lead to a need for more perfect adherence levels to the given therapy. Additionally, Figure 3 implies that the initial viral strain of the patient plays a critical role in the progression of HIV dynamics. According to the Table 3, NRTI therapy combinations yield 38.4% and 78% decrease in infection rate on average (among all therapies) (95% CI [36.2%,40.7%] and [69.7%,86.3%]) for the worst and best case scenario (having most and least resistant initial strain), respectively.

### Details of Model Simulations

In our simulations, we investigate the effect of the type of NRTI therapy, timing of NRTI therapy, and adherence to the given therapy on CD4 + T cell counts of the patients. All possible 21 mono and dual NRTI combinations of six inhibitors have been included in the simulations by considering their independent or additive actions. Initiation time of the NRTI therapy is considered within the first year after the patient became infected and denoted by *τ*. The adherence level of a patient to the given therapy protocol is assigned to a real number *α* between 0 and 1, representing nonadherence to full adherence levels. After initiation of adherence level *α* in a day of the simulation, the patient takes drug(s) with probability *α* according to the parameters given in Table 1. The initial viral load, CD4 + T cell count and macrophage cell count in the simulations are considered as 1 virion/ml, 10^4^ cell/ml and 150 cell/ml, respectively (Hernandez-Vargas, 2019).

**Table 1.** Drug specific parameters for time-dependent drug efficacy equation (4).

| Parameter/Drug | 3TC | ABC | AZT | D4T | DDI | TDF |
| --- | --- | --- | --- | --- | --- | --- |
| $IC_{50}$ ($\times 10^{-5}$ mg/ml) | 3.97 | 132.64 | 1.87 | 4.25 | 113.11 | 16.24 |
| $D$ (mg) | 300 | 300 | 300 | 40 | 400 | 300 |
| $I_d$ (day) | 1 | 0.5 | 0.5 | 0.5 | 1 | 1 |
| $F$ | 0.86 | 0.83 | 0.64 | 0.86 | 0.42 | 0.39 |
| $k_a$ | 27.98 | 51.07 | 37.42 | 54.29 | 32.34 | 8.36 |
| $k_e$ | 3.44 | 8.52 | 14.25 | 7.84 | 47.30 | 16.58 |
| $V_d$ (ml) | 91000 | 60200 | 112000 | 46000 | 54000 | 87500 |

**Table 2.** Parameter values, descriptions, and references of within-host models (1) and (5).

| Parameter | Value | Unit | Reference/ Parameter Variation |
| --- | --- | --- | --- |
| $s_T$ | 10000 | $d^{-1}$ | Perelson and Nelson (1999) |
| $s_M$ | 150 | $d^{-1}$ | Perelson and Nelson (1999) |
| $k_T$ | $4 \times 10^{-8}$ | $mld^{-1}$ | Hadjiandreou et al. (2007)<br>$3.2 \times 10^{-8} - 10^{-7}$ |
| $k_M$ | $4 \times 10^{-11}$ | $mld^{-1}$ | Adjusted according to Hernandez-Vargas (2019) |
| $p_T$ | 38 | $d^{-1}$ | Hernandez-Vargas (2019)<br>30.4-114 |
| $p_M$ | 35 | $d^{-1}$ | Hernandez-Vargas (2019)<br>22-132 |
| $\delta_T$ | 0.01 | $d^{-1}$ | Hadjiandreou et al. (2007)<br>0.001-0.017 |
| $\delta_{T^*}$ | 0.4 | $d^{-1}$ | Perelson and Nelson (1999)<br>0.1-0.45 |
| $\delta_M$ | 0.001 | $d^{-1}$ | Hernandez-Vargas (2019)<br>$10^{-4} - 1.4 \times 10^{-3}$ |
| $\delta_{M^*}$ | 0.001 | $d^{-1}$ | Hernandez-Vargas (2019)<br>$10^{-4} - 1.2 \times 10^{-3}$ |
| $\delta_V$ | 2.4 | $d^{-1}$ | Hadjiandreou et al. (2007)<br>0.96-2.64 |
| $\rho_T$ | 0.0077 | $d^{-1}$ | Adjusted according to Hernandez-Vargas (2019) |
| $\rho_M$ | 0.0025 | $d^{-1}$ | Adjusted according to Hernandez-Vargas (2019) |
| $c_T$ | 300 | $ml^{-1}$ | Adjusted according to Hernandez-Vargas (2019) |
| $c_M$ | 220 | $ml^{-1}$ | Adjusted according to Hernandez-Vargas (2019) |

**Table 3.** Descriptive statistics (×10^−8^) of infection rate *β*_*i*_ values for all possible mono and dual NRTI therapies.

| Drugs | Mean | Max | Min | Std | Median | Mode | $Q_1$ | $Q_3$ |
| --- | --- | --- | --- | --- | --- | --- | --- | --- |
| D4T-3TC | 1.290 | 0.160 | 2.069 | 0.363 | 1.295 | 0.16 | 1.029 | 1.576 |
| D4T-AZT | 1.370 | 0.427 | 2.358 | 0.442 | 1.368 | 0.427 | 1.021 | 1.722 |
| TDF-D4T | 1.403 | 0.604 | 2.373 | 0.371 | 1.382 | 0.604 | 1.109 | 1.679 |
| D4T | 1.442 | 0.697 | 2.319 | 0.351 | 1.426 | 0.697 | 1.157 | 1.72 |
| AZT-3TC | 1.473 | 0.154 | 2.212 | 0.462 | 1.531 | 0.154 | 1.109 | 1.877 |
| D4T-ABC | 1.525 | 0.695 | 2.405 | 0.374 | 1.514 | 0.695 | 1.221 | 1.826 |
| DDI-D4T | 1.592 | 0.765 | 2.466 | 0.382 | 1.581 | 0.765 | 1.279 | 1.903 |
| TDF-AZT | 1.627 | 0.474 | 2.523 | 0.506 | 1.683 | 0.474 | 1.226 | 2.056 |
| AZT-ABC | 1.755 | 0.551 | 2.529 | 0.503 | 1.844 | 0.551 | 1.363 | 2.194 |
| AZT | 1.775 | 0.554 | 2.564 | 0.513 | 1.858 | 0.554 | 1.373 | 2.223 |
| DDI-AZT | 1.834 | 0.562 | 2.602 | 0.533 | 1.933 | 0.562 | 1.412 | 2.307 |
| TDF-3TC | 1.884 | 0.3 | 2.265 | 0.307 | 1.952 | 0.3 | 1.835 | 2.065 |
| 3TC | 1.965 | 0.29 | 2.173 | 0.339 | 2.114 | 0.29 | 2.000 | 2.133 |
| ABC-3TC | 2.030 | 0.274 | 2.318 | 0.359 | 2.173 | 0.274 | 2.025 | 2.225 |
| DDI-3TC | 2.155 | 0.323 | 2.373 | 0.356 | 2.305 | 0.323 | 2.201 | 2.325 |
| TDF-ABC | 2.172 | 1.508 | 2.588 | 0.181 | 2.176 | 1.508 | 2.038 | 2.314 |
| TDF | 2.299 | 1.889 | 2.665 | 0.172 | 2.3 | 1.889 | 2.163 | 2.438 |
| TDF-DDI | 2.323 | 1.917 | 2.667 | 0.164 | 2.327 | 1.917 | 2.194 | 2.457 |
| ABC | 2.459 | 1.733 | 2.698 | 0.126 | 2.485 | 1.733 | 2.404 | 2.544 |
| DDI-ABC | 2.546 | 1.869 | 2.726 | 0.106 | 2.57 | 1.869 | 2.505 | 2.617 |
| DDI | 2.746 | 2.675 | 2.78 | 0.02 | 2.747 | 2.675 | 2.732 | 2.762 |

It is assumed that the patient is infected with one type of mutant strain with one to five-point mutations on the reverse transcriptase enzymes. In this way, five groups are constructed so that each group includes five different strains. These viral strains have been determined according to the frequency of presence in the Stanford HIV drug resistance database. These sets of initial viral strains are denoted by *G*_*ij*_ where *i* = 1, 2, 3, 4, 5 denotes the number of the point mutations in the strain and *j* = 1, 2, 3, 4, 5 indexes the examples that are most frequently occurred in the dataset. We have performed our simulations with these 25 different initial viral strains having the following point mutations: *G*_11_ = {69*D*}, *G*_12_ = {70*E*},*G*_13_ = {74*I*}, *G*_14_ = {151*M*}, *G*_15_ = {41*L*}, *G*_21_ = {69*D*, 115*F*},*G*_22_ = {69*D*, 215*Y*}, *G*_23_ = {70*R*, 215*Y*}, *G*_24_ = {67*N*, 69*D*},*G*_25_ = {67*N*, 70*R*}, *G*_31_ = {69*D*, 115*F*, 215*Y*}, *G*_32_ = {69*D*, 70*R*, 115*F*}, *G*_33_ = {67*N*, 69*D*, 215*Y*}, *G*_34_ = {67*N*, 70*R*, 215*Y*}, *G*_35_ = {67*N*, 69*D*, 70*R*}, *G*_41_ = {67*N*, 69*D*, 115*F*, 215*Y*}, *G*_42_ = {67*N*, 70*R*, 115*F*, 215*Y*}, *G*_43_ = {69*D*, 70*R*, 115*F*, 215*Y*}, *G*_44_ = {67*N*, 69*D*, 70*R*, 115*F*}, *G*_45_ = {65*N*, 69*D*, 70*R*, 215*Y*}, *G*_51_ = {65*N*, 69*D*, 70*R*, 115*F*, 215*Y, G*_52_ = {69*D*, 70*R*, 74*F*, 115*F*, 215*Y*}, *G*_53_ = {41*L*, 67*N*, 69*D*, 70*R*, 215*Y*}, *G*_54_ = {65*N*, 67*N*, 69*D*, 70*R*, 215*Y*}, *G*_55_ = {67*N*, 69*D*, 70*R*, 74*I*, 215*Y*}. For instance, *G*_14_ = {151*M*} strain has only one point mutation 151*M* and the rest of the amino acids are same as for the wild-type.

### Measuring the Therapy Success

It is essential to track the success of the antiretroviral therapy by protecting the patients from the AIDS phase, i.e. by keeping the CD4 + T cell count as high as possible. The AIDS phase yields opportunistic infections for the patients and occurs when CD4 + T cell count is less than 200 cell/*µ*l (Kitahata et al., 2009). Our primary metric for the success of the NRTI therapy is the occurrence and nonoccurrence of the AIDS phase after initiating the therapy with some initiation timing *τ* and adherence level *α* as also done by cohort studies (van Sighem et al., 2003). All simulations start with one infected CD4 + T cell and one infected macrophage cell with one of the initial strains *G*_*ij*_. The simulation final time *t* _*f*_ is considered as 20 years, and the therapy success/failure is determined according to the occurrence of AIDS phase in 20 years. We run our simulations for randomly scattered 512 (*α, τ*) ∈ [0, 1] × [0, 365] pairs for predetermined initial strain *G*_*ij*_. The success rate (SR) of a therapy is measured as the number of (*α, τ*) pairs that lead to protection from AIDS phase in all 512 (*α, τ*) pairs.

In Figure 4, we show some representative simulation results of the multi-strain within host model (5), starting with the *G*_11_ = {69*D*} strain under various mono and dual NRTI therapies with randomly scattered (*α, τ*) pairs. For this simulation setup, 11 out of 21 NRTI therapy protocols have considerable success in preventing the patient from the AIDS phase. The importance of adherence level (*α*) and initiation timing (*τ*) is evident from the figure for all cases. Especially, initiation timing dominantly determines the distribution of failure/success outcomes and higher *τ* values are seen to yield therapy failure even for high adherence levels. As we observe from the figure, the D4T-AZT combination yields the best SR value by performing well for late initiation with perfect adherence levels. For the current case, the success of D4T-AZT combination is mainly because of the behaviours of the therapy at the higher initiation timing (*τ*) region.

**Figure 4.**
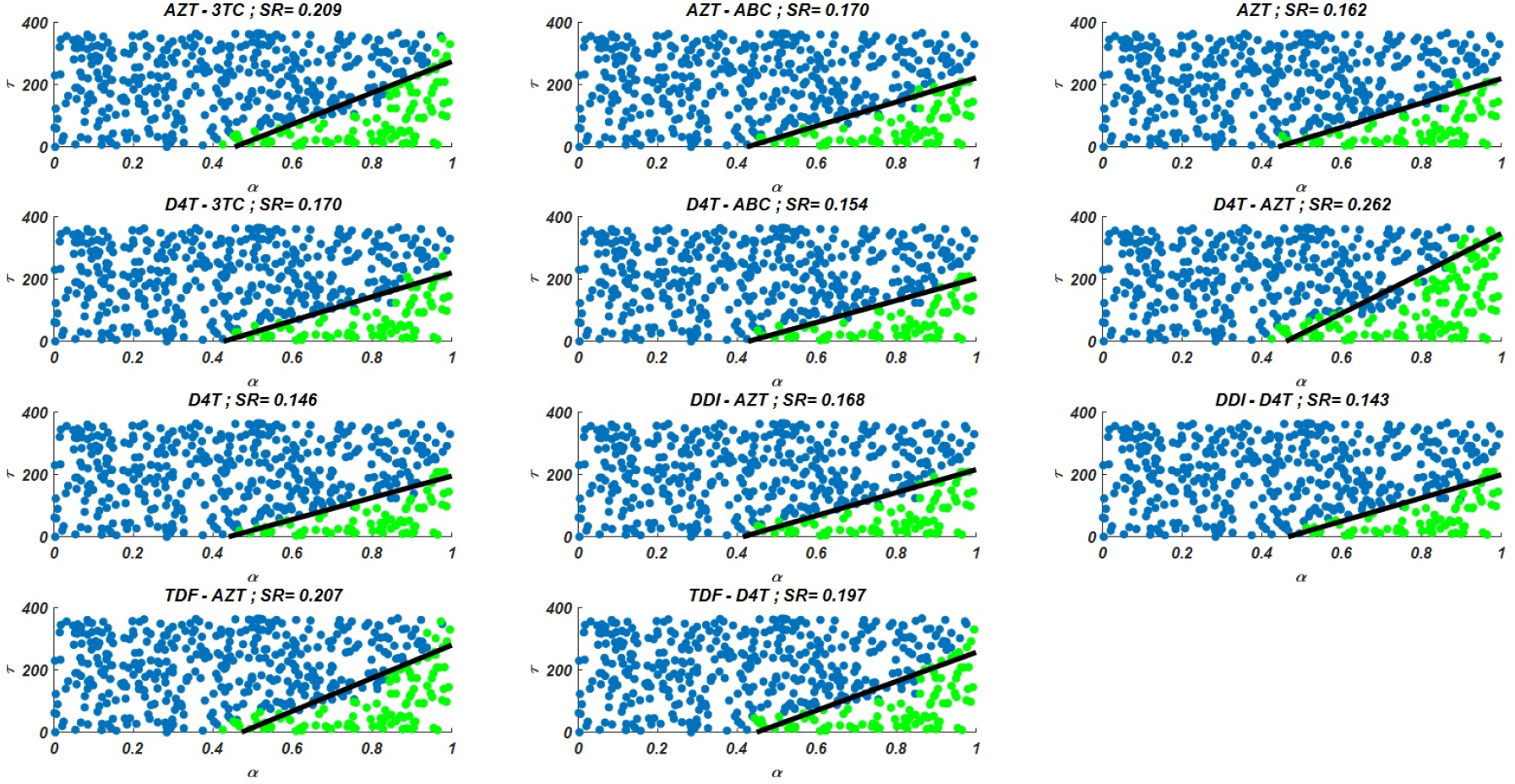
Illustration of possible mono and dual NRTI therapy outcomes carried out using 512 random (*α, τ*) pairs in the current multi-strain within-host model (5). The initial strain has been selected as *G*_11_ = {69*D*}. Blue circles represent the failure after 20 years of simulation, i.e. the AIDS phase occurs when the patients start the therapy *τ* after infection and take the therapy with an adherence rate *α*. Green circles mean that the therapy success under the conditions mentioned above. The SR values represent the success rate defined as SR = # of green circles/# of all circles.

While the importance of the adherence levels is evident from its direct relation with infection rates, the importance of the initiation timing is non-evident and should be explained here clearly. In 4, we illustrate the effect of initiation timing *τ* in our multi-strain model (5) when initial strain and adherence level are selected as *G*_11_ = {69*D*} and *α* = 0.8. According to Figure 5A-B, *τ* = 100 yields successful therapy by maintaining healthy CD4 +T cell and macrophage cells at normal levels and declining the viral load to the undetectable levels. On the other hand, when we assume the initiation timing as *τ* = 300, virologic failure and AIDS phase are observed in Figure 5C-D. According to our model (5), the main difference between early and late initiation timing is the level of virulence and healthy cells at that time. Model implies that higher virulence leads (*V*_*i*_≫1) considerable contribution of healthy CD4 + T cell and macrophage cell proliferation terms (with approximately *ρ*_*T*_ *T* and *ρ*_*M*_*M*). According to the dynamics, in some non-optimal cases (when the infection rate is not considerably reduced), the availability of more healthy cell reservoirs leads to the reoccurrence of the viral strains even under the existence of NRTI therapies. Since the infection rate of virions to macrophage cells is less than CD4 + T cells (*k*_*M*_ *< k*_*T*_), macrophage cells become the main prey of virions in the reoccurrence phase when any non-optimal therapy is implemented. Thus, the level and type of the NRTI therapy should be planned so that the reoccurrence of the viral strains should be blocked depending on the initiation time *τ*. Additionally, in the reoccurrence phase of viral strains, non-perfect adherence to the therapy leads to the selection of resistant strains (Figure 5D). In this case, two possible issues arise:

1. If the therapy protocol of the patient is updated, the probability of therapy success is less than the first time he/she starts the therapy.
2. The probability of infecting another person with more resistant strains increases, and the probability of having an AIDS phase increases for the infected person.

The other difference between early and late initiations is the occurrence of new drug-resistant strains before treatment without selection. These new strains may contribute to the failure when late initiation happens. In some scenarios, in addition to the effect of more healthy cells on the reoccurrence of the viral strains, these mutated strains may lead to virologic failure and the occurrence of the AIDS phase. Since we assume that all viral strains have equal fitness costs, only one-point mutants of initial strains can be seen in the host before treatment. However, the existence of low viral loads of new mutated strains is enough for selecting these strains after antiretroviral therapy. Additionally, if the fitness costs of viral strains were available and these values were used in the model (5), the contribution of these pre-treatment mutations to the emergence of the AIDS phase would be more than the current case. Thus, according to our simulations, initiation timing is as crucial as the adherence level to overcome the AIDS phase and to protect the possible susceptible persons from more dangerous scenarios.

**Figure 5.**
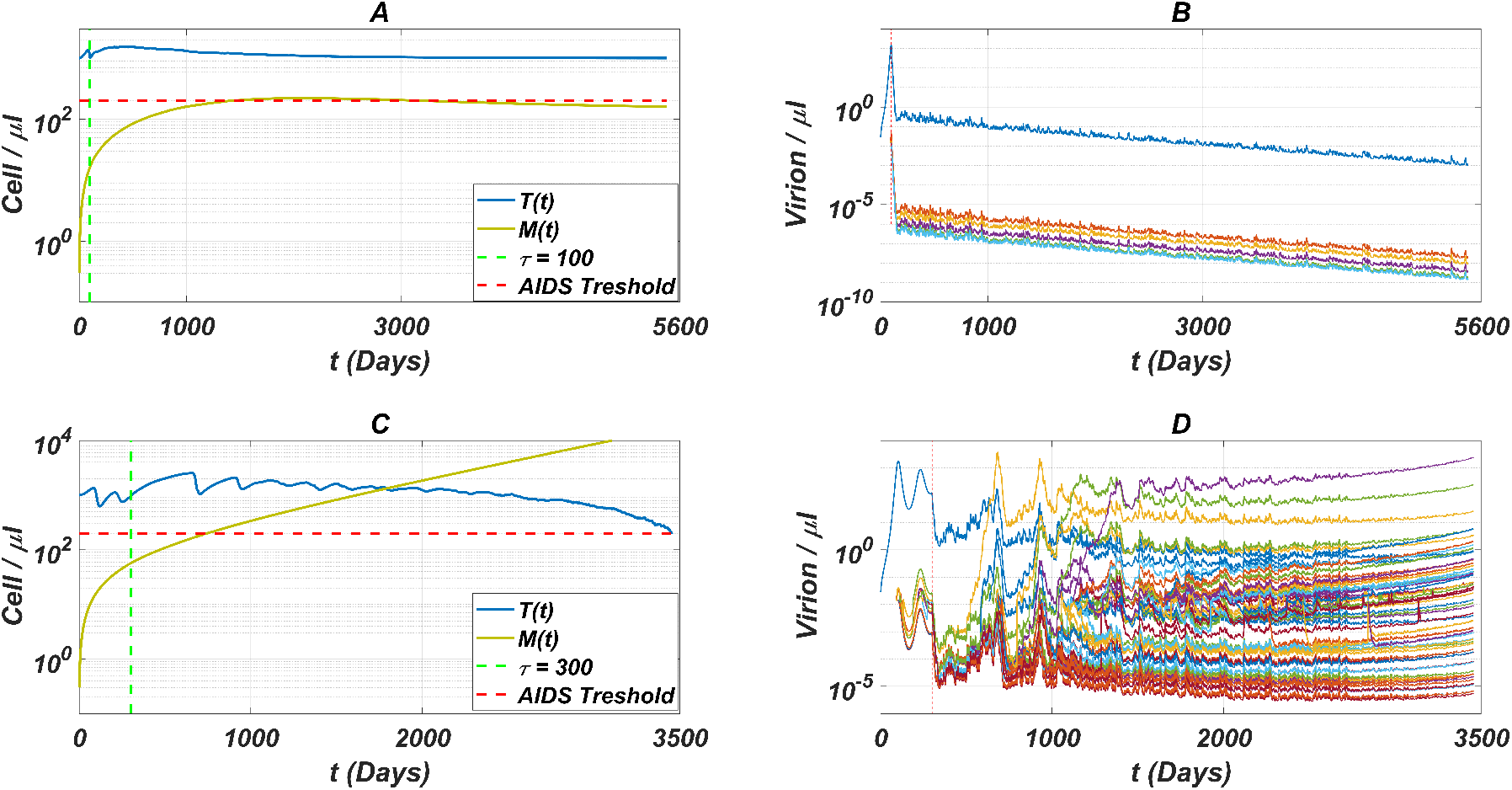
The effect of initiation timing is illustrated with healthy cell and virion counts. The initial strain is taken as *G*_11_ = {69*D*} and the common adherence level *α* = 0.8 is considered. A) Dynamics of T(t) and M(t) when *τ* = 100, B) Dynamics of viral strains when *τ* = 100, C) Dynamics of T(t) and M(t) when *τ* = 300, D) Dynamics of viral strains when *τ* = 300.

### General Simulation Results

Here we have simulated our multi-strain within-host model (5) for all possible initial strains *G*_*ij*_ to observe the effect of initial strains on the success rates. All possible mono and dual NRTI therapies have been implemented for randomly scattered 512 (*α, τ*) ∈ [0, 1] × [0, 365] pairs. The SR values of mono and dual NRTI therapies are calculated, and the well-performed combination results are comparatively illustrated in Figure 6.

**Figure 6.**
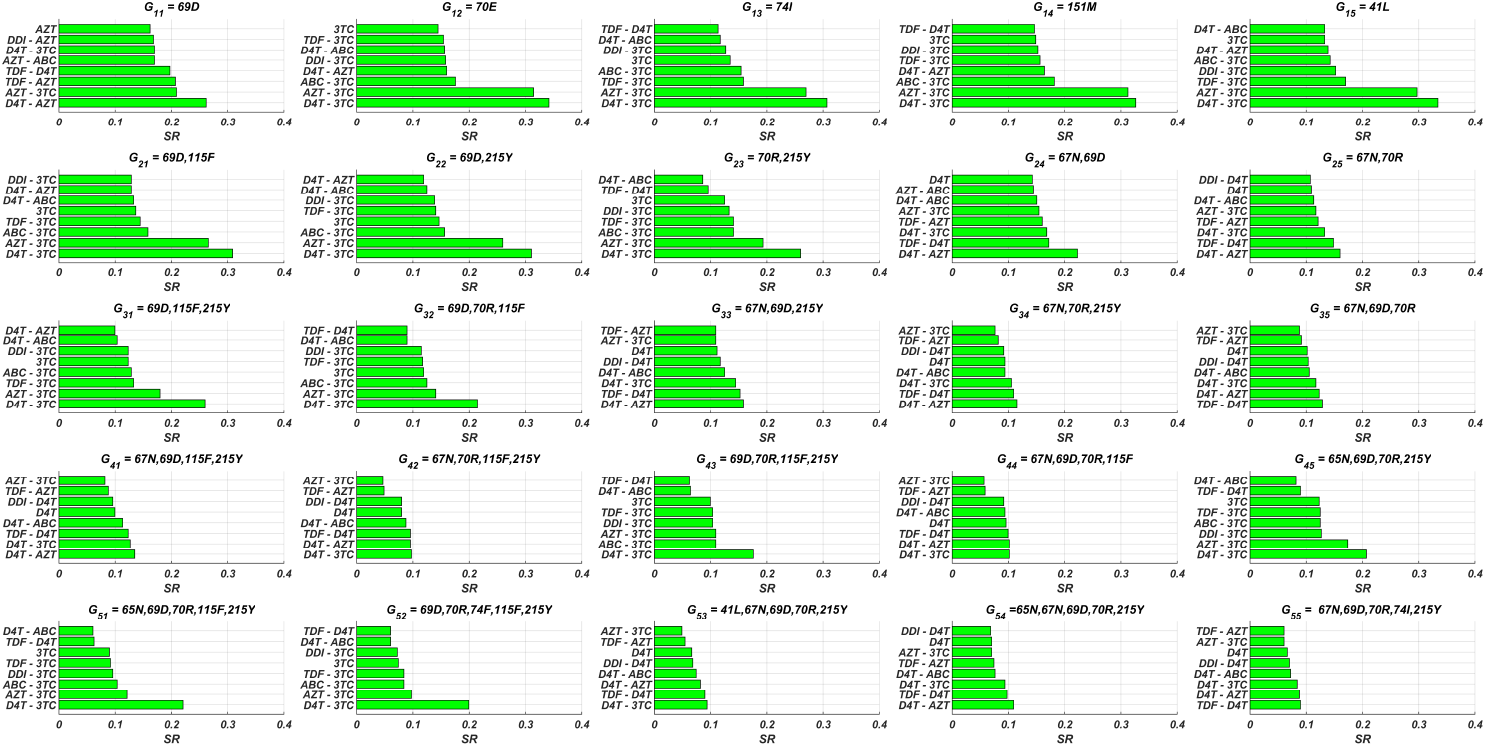
SR values of various NRTI combinations obtained by simulating multi-strain within host model (5) with initial viral strain *G*_*ij*_ for randomly scattered 512 (*α, τ*) ∈ [0, 1] × [0, 365] pairs.

In line with Figure 3 and Table 3, the D4T-3TC combination has been the best option for 16 out of 25 cases. The inhibitory potential of this combination is because of the pharmacokinetic parameters (see Table 1) of inhibitors, the drug-resistance profiles of inhibitors (see Table 3) and their Bliss independent action on the target enzyme. Following the D4T-3TC combination, D4T-AZT and TDF-D4T combinations are observed to be in first place in 7 and 2 out of 25 cases, respectively. The strong relation between the infection rate of an initial strain (and possible new strains) and the corresponding success rate value is evident from the correlation between Figures 3 and 6.

According to the our modelling framework, since the fitness cost of all strains are assumed to be the same, the initial strains become dominant when the patient is diagnosed to start an NRTI therapy. As evident also from Figure 5D, mutational variations are less likely to be detected (less than 0.05 copies/*µ*l) when a phenosense assay is implemented. Thus, the clinician would observe the initial strain as well as a few mutational variations to make a decision about the NRTI therapy protocol. It is inevitable to ask whether the only predictor of the success rate is the infection rate of the initial strain. Our alternative hypothesis is that the existing mutational variations (detected or undetected one-point mutants of the initial strain) before the treatment also have a considerable effect on the success rates.

We have constructed two artificial neural network (ANN) models to determine the more accurate predictors of the success rates (see Figure 7 for model architectures). In the first model (Model 1), the ANN function with one hidden layer and six neurons maps the infection rate of the initial strain to the success rate. In the second one (Model 2), the ANN function with the same architecture maps (*x, y, z*) triple to success rate where *x* is the infection rate of the initial strain, *y* and *z* are the mean and maximum values of infection rates among all possible neighbour mutants of the initial strain. Thus, Model 1 hypothesizes that the only predictor of the success rate is the infection rate of the initial strain. In contrast, Model 2 hypothesizes that both the infection rates of the initial strain and its neighbour strains affect the success rate. Simulation results are given in Figure 6 for 25 initial strains converted to the training data for these machine learning models. 304 input-output relations have been obtained, and this data is divided into the training, testing, and validation sets (70%, 15% and 15%). Using the scaled conjugate gradient algorithm, these two models are trained, and the regression performances of both models are given in Figures 8-9.

**Figure 7.**
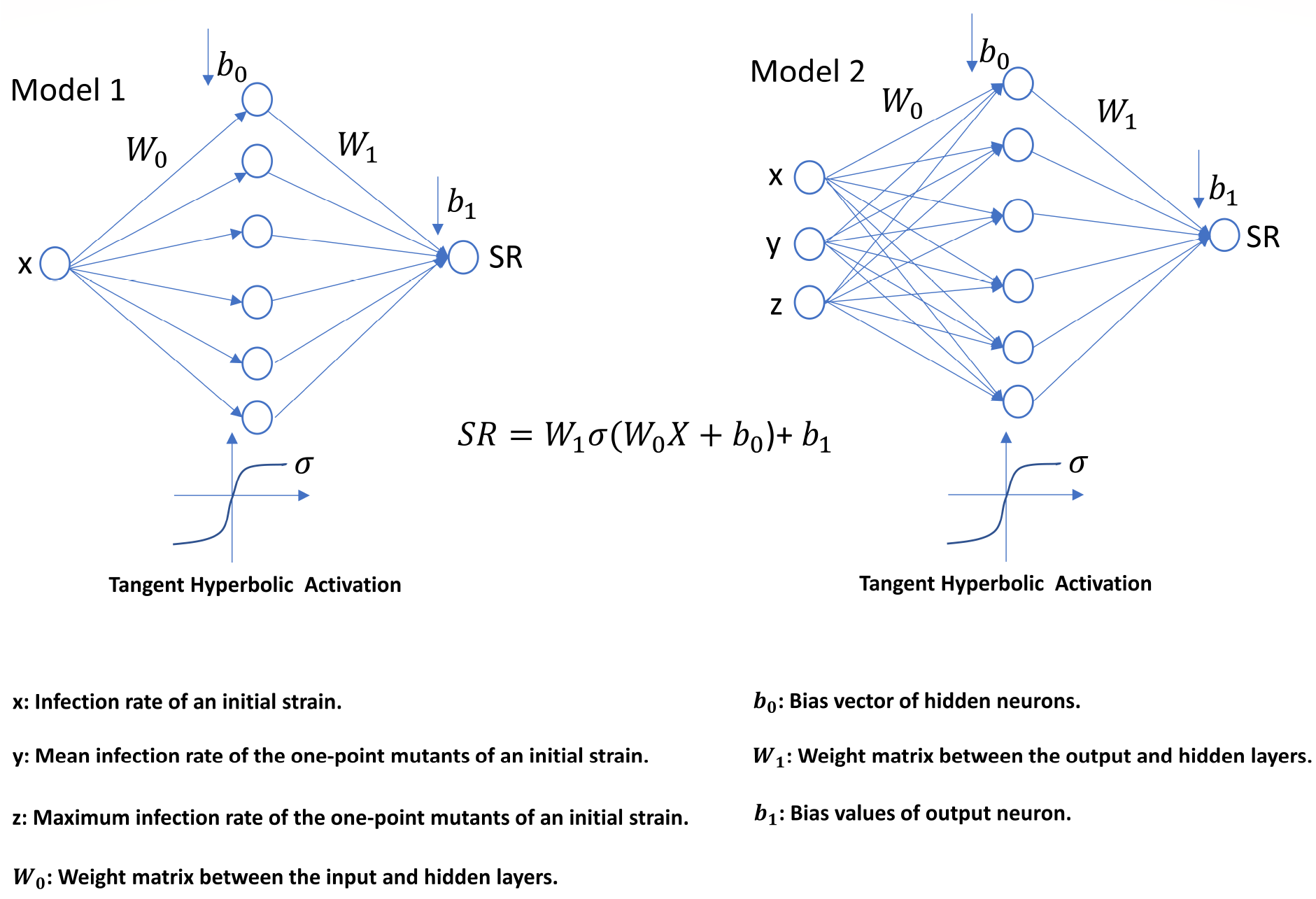
Artificial neural network architectures of Model 1 and Model 2.

**Figure 8.**
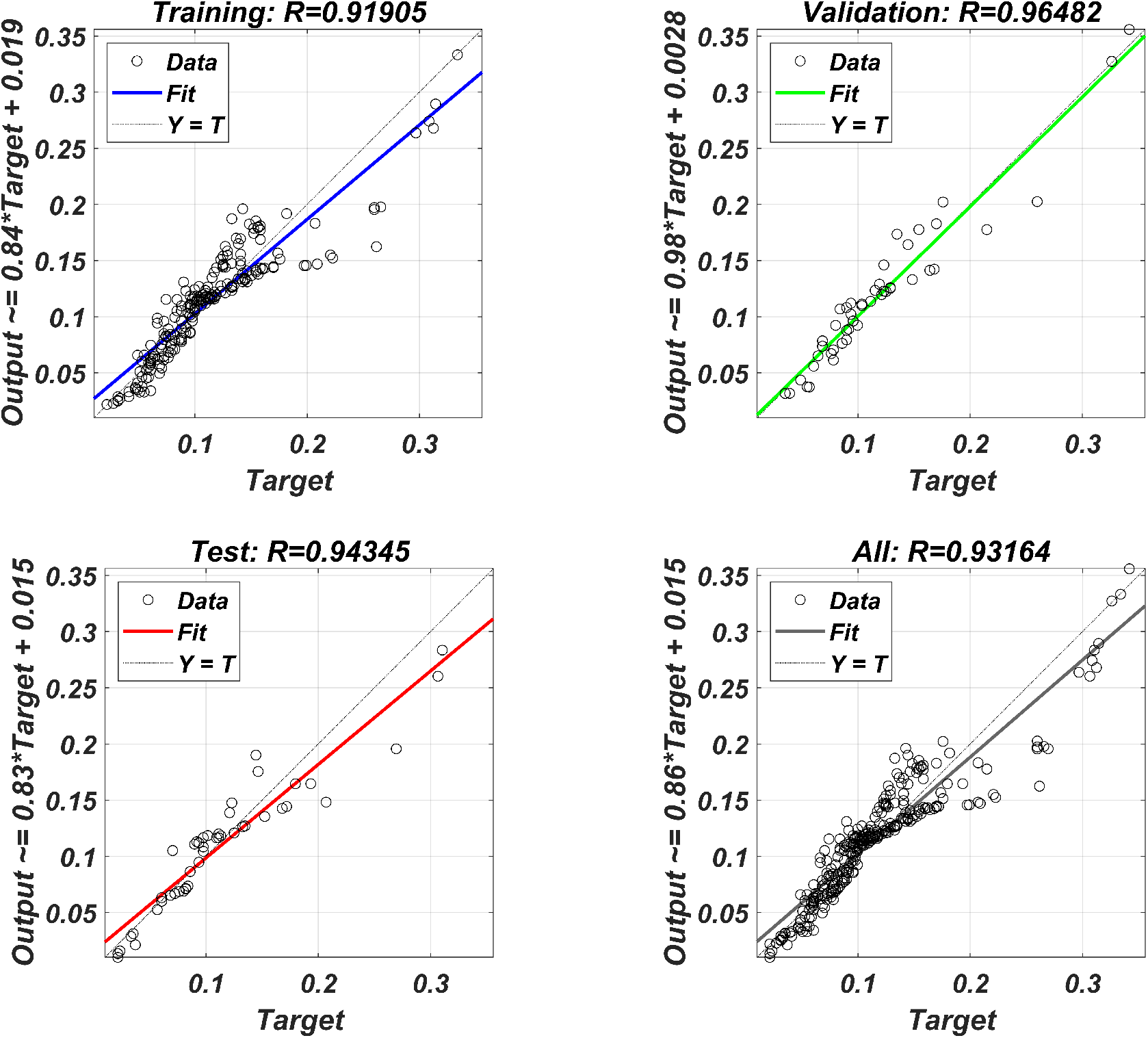
Regression performances of Model 1 predicting the success rates from the infection rates of the initial strains.

**Figure 9.**
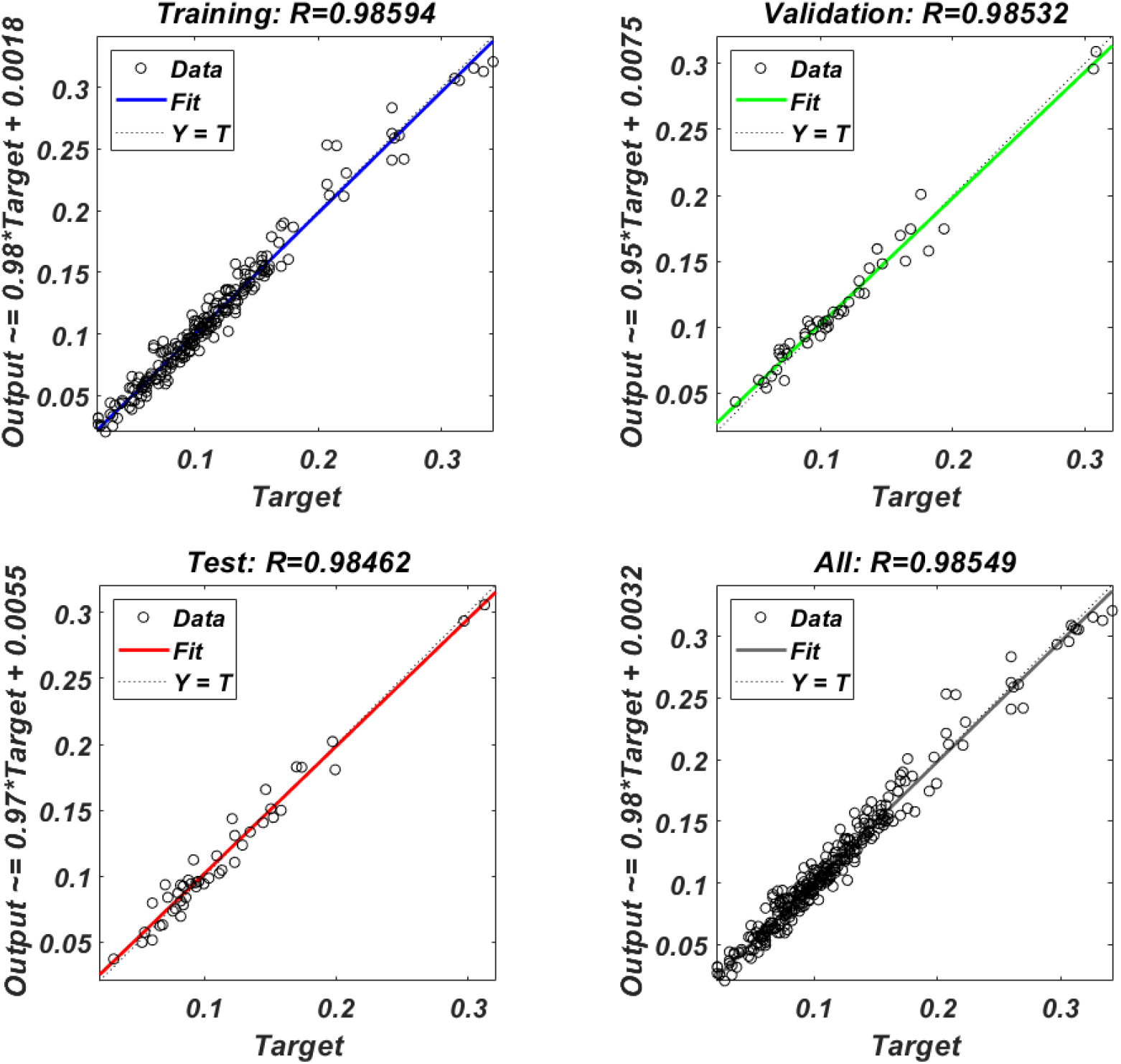
Regression performances of Model 2 predicting the success rates from the infection rates of both initial strains and one-point mutant strains.

Figures 8-9 imply that predicting the success rates from the infection rates with Models 1-2 have excellent accuracy on the internal training, test, and validation data sets. Model 2 performs better regression performances than Model 1 on the internal data set generated from the simulation results illustrated in Figure 6. To realistically test the prediction accuracies of these machine learning models, we have generated external test data set by simulating the model (5) with 25 random initial strains having one-to-five-point mutations. Regression performances of both the Model 1 and Model 2 on this external test set are illustrated in Figure 10. The figure shows that Model 2 is far more accurate than Model 1. This observation indicates that the success rate of a therapy combination should be predicted by not only considering the infection rates of the initial strains but also the infection rates of possible one-point mutant strains (detected or undetected during phenosense assays). These mutant strains play a vital around the boundary line of the success/failure distribution on the (*τ, α*) plane (for instance, see Figure 4). Mutant strains may occur in the host without being dominant, and they may be selected after initiation of the antiretroviral therapy and this affects the success/failure outcome.

**Figure 10.**
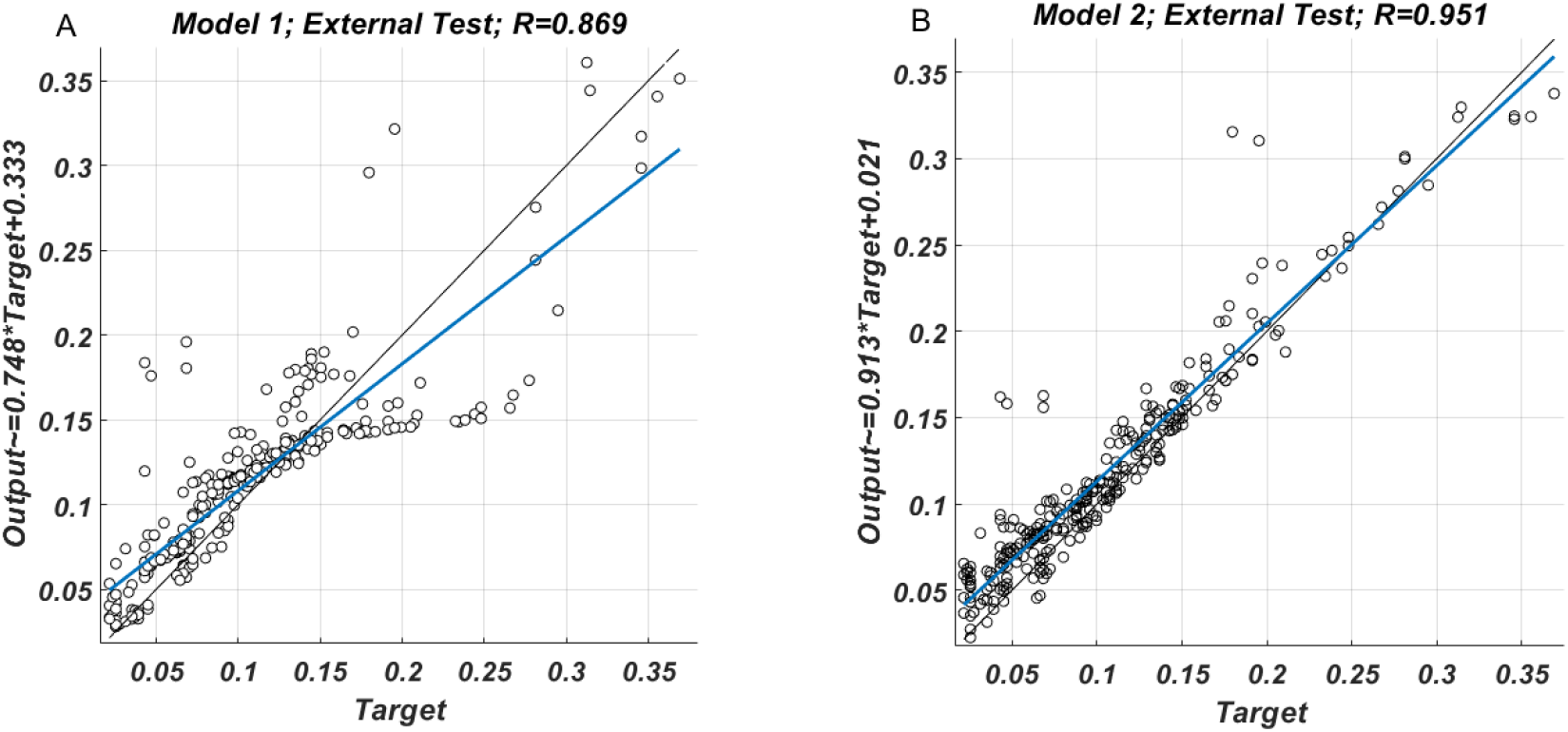
Regression performances of the Model 1 (A) and Model 2 (B) on the external test data set.

## DISCUSSIONS AND CONCLUSIONS

In this study, we have proposed a multi-strain within-host model of HIV infection with time-dependent NRTI therapy. Drug-resistant strains have been assumed to initiate the infection for the patients, and six available NRTI inhibitors with mono and dual combinations have been implemented in the simulations for various initiation timing and adherence levels. In order to assess the drug response curves with IC50 values of the NRTI resistant strains, artificial neural network models are trained for each inhibitor by using the Stanford HIV drug resistance database. To elucidate the time-drug efficiency and time-infection rate curves, the pharmacokinetic parameters of the inhibitors have been computed and hybridized with the corresponding IC50 values. We have designed our simulation environment to determine the impact of initial strains, initiation timing for the therapy protocol and adherence levels to the given drug usage schedule on the occurrence of AIDS phase within 20 years after infection.

Based on our modelling framework, the occurrence of the AIDS phase has appeared to be highly correlated with the initiation timing and adherence level of NRTI therapies. The success rate of the NRTI therapies in case of late initiation has led to the availability of more macrophage reservoirs for viral strains and emergence of more resistant strains, and then reoccurrence of resistant strains after an initial decline. Although some mathematical models assume implicitly that the initiation timing does not affect the success-failure of the therapy (Dixit and Perelson, 2004; Rong et al., 2007), our multi-strain model more realistically catches the penalty of late initiation since the late initiation was proven to block the therapy success in various experimental results (Kitahata et al., 2009; van Sighem et al., 2003). Our simulation results have revealed that in the case of the late initiation to therapy, the efficiency of the therapy should be much greater than the early initiation case in order to prevent the possible AIDS phase.

We have shown that D4T-3TC, D4T-AZT and TDF-D4T combinations are more likely to prevent the patients from the AIDS phase for various initial strains that are the most common ones in the Stanford HIV drug resistance database. It has been observed that these inhibitors provide less infection rates in the presence of various viral strains due to their pharmacokinetic parameters and IC50 values. Based on our machine learning predictions of infection rates and simulation results, it has been proven that the success rate of a therapy for an initial strain is accurately predicted by the infection rates of both the first strains and possible mutants of the first strains. This observation implies that the emergence of some neighbor strains that mutated from an initial strain is likely to have a considerable effect on the success/failure outcome of the therapy. Therefore, considering the detected viral strain and possible detect/undetected neighbouring mutant strains, it is more reasonable to recommend the optimal therapy combinations to patients.

An inevitable criticism of our model is the assumption of a unique fitness cost for each mutant strain. It has been explained that this assumption is due to the lack of experimental fitness costs of each possible strain. Additionally, according to our research, there is not much data for specific strains to construct a machine learning model as we did for IC50 values. The theoretical perspective shows that the cost of resistance plays a vital role in the selection of resistant strains and and the strain-dependent fitness costs would change our success rate values positively or negatively depending on the scenario. In addition, success/failure rates will be more affected by the infection rates of possible mutant strains than the current case, as the strain-dependent fitness costs may lead to selection of neighbouring strains before the initiation of the therapy. Therefore, the availability of fitness cost data for various NRTI-related strains will increase the reliability of our estimates of success rates under our simulation framework.

In this study, the effect of NRTI inhibitors, which are the most important members of Highly Active Antiretroviral Therapy (HAART) (Achhra and Boyd, 2013), on the dynamics of healthy cells and viral strains has been investigated. As the Stanford drug resistance database also includes genotype-phenotype data for protease inhibitors (PI), non-nucleotide reverse transcriptase inhibitors (NNRTI), and integrase inhibitors (II), some future studies may include these groups of inhibitors with possible mono, dual or triple drug combinations. Some existing HAART protocols may also be simulated through such a modelling framework. On the other hand, we did not consider the too late initiation of the NRTI therapy at very low CD4 + T cell levels, due to the failure of currently available therapy protocols in such cases. Some future studies may also concentrate on the investigation of the required therapies, including NRTI, PI, NNRTI, or II inhibitors, to protect patients from the AIDS stage when the patient is diagnosed too late.

## Data Availability

All data produced are available online at https://hivdb.stanford.edu/download/GenoPhenoDatasets/PI_DataSet.txt.

## CODE AND SOFTWARE AVAILABILITY

All data and necessary codes are deposited to: https://github.com/tnchsyn/multistrainhivmodel.

