## Supplementary Material for "Machine learning aided multiscale modelling of the HIV-1 infection in the presence of NRTI therapy"

### Supplementary Article

According to the Dixit and Perelson (2004), single-dose version of equation (4) can be expressed as follows

$$C_b(t) = \frac{FDk_a}{V_d(k_e - k_a)}[e^{-k_a t} - e^{-k_e t}]. \quad \text{(SM1)}$$

The well known drug-specific parameters $C_{max}$ (maximum concentration of a drug in the blood) $AUC$ (area under the concentration-time curve) and $t_{max}$ (the at which $C_{max}$ occurs) can be derived from equation (SM1) as

$$C_{max} = \frac{FD}{V_d}\left(\frac{k_e}{k_a}\right)^{\frac{k_e}{k_e - k_a}}, \quad \text{(SM2)}$$

$$AUC = \int_0^\infty C_b(t)dt = \frac{FD}{V_d k_e}, \quad \text{(SM3)}$$

$$t_{max} = \frac{ln\left(\frac{k_e}{k_a}\right)}{k_a - k_e}. \quad \text{(SM4)}$$

$C_{max}$, $AUC$ and $t_{max}$ values are available for all NRTI drugs, and estimation of the pharmacokinetic parameters $k_a$ and $k_e$ can be done by solving equations (SM2)-(SM4) in the least-squares sense. In this way, we have evaluated $k_a$ and $k_e$ values of all drugs with the use of experimental $C_{max}$, $AUC$ and $t_{max}$ presented in Table S1 with their sources.

### Supplementary Table

Table S1. Pharmacokinetic parameters of nucleotide reverse transcriptase inhibitors.

| Parameter/Drug | 3TC | ABC | AZT | D4T | DDI | TDF |
|---|---|---|---|---|---|---|
| $IC_{50}$ Range ($\times 10^{-5}$ mg/ml) | [0.04-343.89] | [105.94-166.07] | [0.26-13.09] | [0.20-89.68] | [56.55-226.23] | [1.14-229.77] |
| $IC_{50}$ Geometric Mean ($\times 10^{-5}$ mg/ml) | 3.97 | 132.64 | 1.87 | 4.25 | 113.11 | 16.24 |
| $D$ (mg) | 300 | 300 | 300 | 40 | 400 | 300 |
| $I_d$ (day) | 1 | 0.5 | 0.5 | 0.5 | 1 | 1 |
| $F$ | 0.86 | 0.83 | 0.64 | 0.86 | 0.42 | 0.39 |
| $k_a$ (Estimated) | 27.98 | 51.07 | 37.42 | 54.29 | 32.34 | 8.36 |
| $k_e$ (Estimated) | 3.44 | 8.52 | 14.25 | 7.84 | 47.30 | 16.58 |
| $V_d$ (ml) | 91000 | 60200 | 112000 | 46000 | 54000 | 87500 |
| Reference | https://www.accessdata.fda.gov/ | https://www.accessdata.fda.gov/ | https://www.accessdata.fda.gov/ | https://www.accessdata.fda.gov/ | https://www.hiv-druginteractions.org/ | https://www.accessdata.fda.gov/ |
